# Daily consumption of ketone ester, bis-octanoyl (R)-1,3-butanediol, is safe and tolerable in healthy older adults, a randomized, parallel arm, double-blind, placebo-controlled, pilot study

**DOI:** 10.1101/2024.05.03.24306699

**Authors:** Brianna J. Stubbs, Elizabeth B. Stephens, Chatura Senadheera, Sawyer Peralta, Stephanie Roa-Diaz, Laura Alexander, Wendie Silverman-Martin, Thelma Y. Garcia, Michi Yukawa, Jenifer Morris, Traci M. Blonquist, James B. Johnson, John C. Newman

## Abstract

**Objectives:** Ketone bodies are endogenous metabolites produced during fasting or a ketogenic diet that have pleiotropic effects on aging pathways. Ketone esters (KEs) are compounds that induce ketosis without dietary changes, but KEs have not been studied in an older adult population. The primary objective of this trial was to determine tolerability and safety of KE ingestion in older adults.

**Design:** Randomized, placebo-controlled, double-blinded, parallel-arm trial, with a 12-week intervention period (**NCT05585762**).

**Setting:** General community, Northern California, USA.

**Participants:** Community-dwelling older adults, independent in activities of daily living, with no unstable acute medical conditions (n=30) were randomized and n=23 (M= 14, F=9) completed the protocol.

**Intervention:** Participants were randomly allocated to consume either KE (bis-octanoyl (R)-1,3-butanediol) or a taste, appearance, and calorie-matched placebo (PLA) containing canola oil.

**Measurements:** Tolerability was assessed using a composite score from a daily log for 2-weeks, and then via a bi-weekly phone interview. Safety was assessed by vital signs and lab tests at screening and weeks 0, 4 and 12, along with tabulation of adverse events.

**Results:** There was no difference in the prespecified primary outcome of proportion of participants reporting moderate or severe nausea, headache, or dizziness on more than one day in a two-week reporting period (KE n =2 (14.3% [90% CI = 2.6 – 38.5]); PLA n=1 (7.1% [90% CI = 0.4 – 29.7]). Dropouts numbered four in the PLA group and two in the KE group. A greater number of symptoms were reported in both groups during the first two weeks; symptoms were reported less frequently between 2 – 12 weeks. There were no clinically relevant changes in safety labs or vital signs in either group.

**Conclusions:** This KE was safe and well-tolerated in healthy older adults. These results provide a foundation for use of KEs in aging research.

**Highlights:**

- Ketones esters induce ketosis without dietary changes and may target aging biology
- Studies of ketone esters were limited in duration and focused on younger adults
- We found ketone esters were safe and tolerable for 12 weeks in healthy older adults

## 1.0 Introduction

Nutrition and aging have long been intertwined through years of research of the effects of dietary restriction and fasting on aging in model organisms [1] and now humans (e.g. CALERIE [2], HALLO-P (NCT05424042)). A deepening understanding of the molecular mechanisms involved in fasting and dietary restriction has led to small molecule interventions now under investigation in clinical trials targeting various aging phenotypes, as well as a popular upswell of public and scientific interest in these dietary strategies as geroscience interventions. Geroscience is a research paradigm based on addressing the biology of aging and are-related disease. The geroscience approach of modulating fundamental aging mechanisms holds promise for generating new therapeutics for multifactorial conditions of aging which contribute to functional decline, disability, and loss of independence in older adults [3, 4].

A key shared element of the metabolic state induced by fasting and dietary restriction is the production and oxidation of ketone bodies. Ketone bodies are small molecules synthesized from lipids primarily in the liver, which circulate in the blood to provide non-glucose energy to extrahepatic tissues. The most abundant circulating ketone body is beta-hydroxybutyrate (BHB). Ketone bodies are synthesized constitutively at low concentrations, but are also strongly induced during fasting [5]; during a prolonged fast, humans can produce up to ∼150 g of ketone bodies per day [6]. The energetic and molecular signaling activities of ketone bodies, including supporting mitochondrial function and regulating inflammatory activation, support a mechanistic role in modulating aging [7]. We, and others, previously demonstrated that implementing a ketogenic diet, a nutritional method characterized by reduced carbohydrate intake and increased fat intake without fasting, extended the healthy lifespan of wild-type mice by slowing declines in overall aging-related functions, murine clinical frailty scores, and muscle strength [8, 9].

Exogenous ketones, including KEs, such as bis-octanoyl-(R)1,3-butanediol (BO-BD), are small molecules that nutritionally deliver ketone bodies without other dietary changes [10, 11]. KEs are hydrolyzed in the gut to release ketogenic fatty acids and a ketogenic alcohol, which are then metabolized in the liver, independent of insulin levels or fasting state, to release ketone bodies [12]. In preclinical models, ketone bodies and KEs have been found to impact multiple aging-relevant outcomes in the muscle [13, 14], heart [15, 16], and immune system [17, 18]. Clinically, KEs have been shown to impact a range of endpoints in younger adults, such as blood glucose control [19–22], physical [23–25], cognitive [26–28], immune [29], and cardiovascular [28, 30] function. These examples support our central hypothesis that ketone bodies delivered through KEs may be a candidate geroscience intervention, to target clinical syndromes of aging via direct action on aging biology. However, studies of KE have been limited in duration (28 days) and focused on younger adults; no KE has been studied in an exclusively older adult population.

To be a feasible geroscience intervention, KE must be both tolerable and safe for daily, prolonged use by older adults. Older adults are typically considered to be a more vulnerable population, with a two-fold higher risk of hospitalization with adverse reactions to interventions compared to younger counterparts [31]. One-month studies of KE in younger adults have consistently demonstrated safety and generally good tolerance, although side effects such as nausea, headache, upper abdominal pain, and diarrhea were occasionally reported at mild intensity [32, 33]. Similarly, a 28-day study of bis hexanoyl (R)-1,3-butanediol (BH-BD), a KE that is closely related to BO-BD, reported an increased proportion of young adult participants who experienced headache, nausea, and dizziness at least once in the BH-BD group, but the absolute incidence was low, and severity was generally mild [34]. The lack of data on KE use in older adults represents a major knowledge gap and barrier to translation in this field.

To facilitate future aging-focused investigations using KE, we undertook this 12-week randomized, placebo-controlled, double-blinded, parallel arm, pilot clinical trial with the objective of generating the first human safety and tolerance data for KEs in a generalizable population of older adults, as well as the longest KE intervention study known to date. Based on the data with younger adults, we hypothesized that the KE would be safe and tolerable in healthy older adults.

## 2.0 Methods

### 2.1 Study Design

The Buck Institute Ketone Ester (BIKE) Study was a randomized, double-blind, placebo-controlled, parallel arm, pilot trial designed to assess the feasibility, safety, and tolerability of up to 25 g/day of KE ingestion for 12 weeks compared to a placebo in older adults, 65 years of age and older (**Figure 1**), in preparation for a planned study of KE in pre-frail older adults (NIH funding ID: **1R01AG081226-01)**. Volunteers underwent consent and screening (Visit 1), and then were confirmed to be eligible for participation by medical officers. Upon confirmation of eligibility, acute ketone kinetics were measured (Visit 2, results described in [35]). Baseline safety measures were collected at day 0 (Visit 3), and daily KE or placebo consumption began at day 1. For 12 weeks (or until Visit 5), participants consumed one daily serving of BO-BD or a visually, calorically and taste-matched placebo that contained a matched mass of non-ketogenic fat (canola oil) formulated into a 75 mL beverage. Participants compelted a Beverage Tolerability Questionnaire (BTQ) after consuming the study beverage each day for the first 14 days (acclimation period) to assess the tolerance of the study beverage. Tolerance was further assessed by verbal administration of the BTQ during phone calls every 2 weeks, starting at week 2. An interim safety assessment was performed at week 4 (Visit 4). Upon completion of the 12 weeks, post-intervention safety measures were collected, without study product consumption on the day of the test visit (Visit 5). An optional kinetics visit took place at least 1 week after Visit 5, measuring acute ketone kinetics after consumption of an equivalent serving of a newly developed powder formulation of BO-BD. This study, and all study material was approved by Advarra IRB (WIRB-Copernicus Group, Puyallup, WA, ID# 20213716, current version 1.3 approved on 16^th^ October 2023) and was conducted in accordance with the Declaration of Helsinki. Signed informed consent was provided by the participants prior to implementing any protocol-specific procedures. The study was registered in the clinicaltrials.gov database: **NCT05585762**. Study visits took place at Buck Institute for Research on Aging (Novato, CA, USA) between November 2022 - January 2024; the trial ended when the last enrolled subject completed the protocol. Details of investigator and institutional conflict of interest management plans can be found in our protocol paper [36].

**Figure 1:**
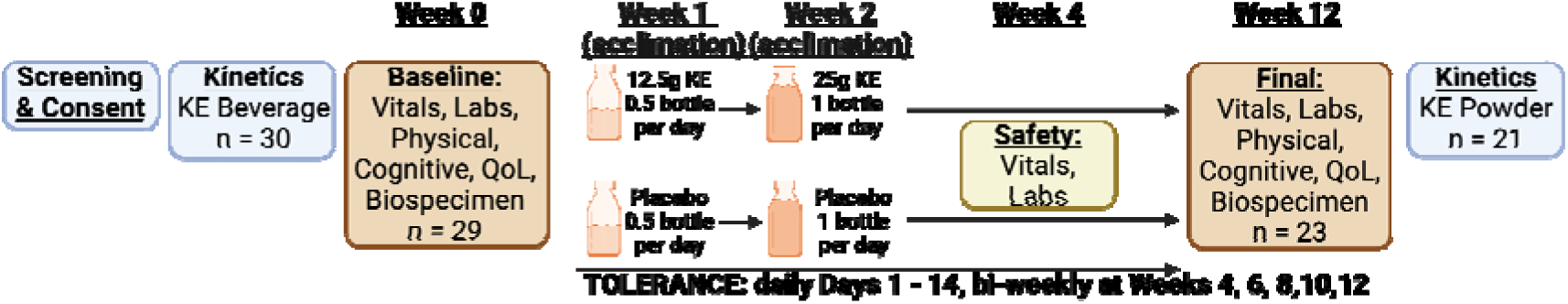
Study Schematic.

### 2.2 Participants and Screening

Participants (n = 30) were community dwelling older adults (≥ 65 years of age) in stable health. We aimed to enroll a broad sample of older adults, allowing common chronic diseases and multiple medications to reflect the intended future study population. Specific exclusion items were chosen based on possible interaction with anticipated KE side effects, such as gastrointestinal (GI) upset. The full inclusion and exclusion criteria have been previously described in our methods paper [36]. At the screening visit, participants completed a medical history questionnaire in addition to assessment of height, weight, body mass index (BMI), vital signs, last menses (females only), current medication / supplement use, and review of inclusion / exclusion criteria to determine eligibility. Additionally, blood and urine were collected for analysis. This information was reviewed by a medical officer for approval to participate. Participants had the opportunity to taste the study beverages to evaluate palatability. Following medical officer approval, participants were randomized during Visit 2 based on order of enrollment to one of the study groups, based on a statistician-generated block allocation sequence (block size 4, intended to equally randomize male and feamle participants). Participants were asked to maintain habitual exercise, meal / diet, and medication / supplement use during the study. The flow of study participants is illustrated in the CONSORT diagram (**Supplementary Figure 1**). Based on studies of symptoms in older adults [37, 38], we predicted the primary outcome rate in the placebo group will be approximately 10%. N=30 (15 per arm) provides 36% power to detect a 25% increase (from 10% to 35%) in the proportion of subjects meeting this primary outcome in the KE condition with two-sided α=0.10. If we assume a larger effect of the KE intervention, N = 30 (15 per arm) provides approximately 75% power to detect an increase (from 10% to 55%) in the proportion of subjects meeting this primary outcome in the KE condition, with two-sided α=0.10.

### 2.3 Study Beverages

The study KE beverage was a tropical-flavored beverage containing the KE bis-octanoyl (R)-1,3-butanediol (Cognitive Switch, BHB Therapeutics Ltd, Dublin, IRE). Each bottle was 75 mL and contained 25 g of KE. Participants were instructed to consume the study beverage at home daily for 12 weeks within 5 minutes of their first meal of the day. Half of a bottle (12.5 g KE) was consumed daily during week 1, and a full bottle (25 g KE) was consumed daily for the remainder of the study. The placebo used in the study was custom manufactured by BHB Therapeutics; the KE was replaced by non-ketogenic canola oil. It was matched for volume, appearance, flavor, and calories. Nutritional facts for study beverages are shown in **Supplementary Table 1**. Beverages were provided as single serving bottles, labeled with the coded group allocation. All personnel involved with the data collection, analysis, and interpretation were blinded to the intervention assigned to participants. Compliance was assessed at week 4 (Visit 4) and week 12 (Visit 5) by asking participants to return any unused product and by review of a daily Study Beverage Log, which was compared to the number of unused bottles returned. Participants who consumed between 80 – 120% of their allocated study product were considered compliant.

### 2.4 Study Questionaires

#### 2.4.1 Study Beverage Log

The daily Study Beverage Log queried compliance with daily beverage intake. Participants noted if they had consumed the beverage, the time of consumption, and noted if they had experienced any tolerance issues or other adverse events (AE).

#### 2.4.2 Beverage Tolerability Questionnaire (BTQ)

The BTQ used in this study was similar to that used in previous tolerability studies [34, 39, 40]. Ten tolerability issues were included in the BTQ: gas / flatulence, nausea, vomiting, abdominal cramping, stomach rumbling, burping, reflux (heartburn), diarrhea, headache, and dizziness. Participants were either asked if the issue occurred since they took the study beverage (paper questionnaire completed daily 3-6 h after KE consumption during 2-week acclimation period) or during the last two weeks (completed verbally during bi-weekly telephone call), and asked to rank the intensity as none, mild (awareness of symptoms but easily tolerated), moderate (discomfort enough to interfere with but not prevent daily activity) or severe (unable to perform usual activity). These correspond to scores of 0-3 respectively for each issue, giving a maximal composite score of 30. After acclimation, participants were asked to report the number of days that each symptom occurred at the respective severity.

### 2.5 In-Clinic Procedures

For testing visits on Days 0, ∼28, and ∼84 (Visits 3, 4, and 5, respectively), participants reported to the clinic after an overnight fast and having avoided exercise, alcohol, and psychoactive cannabis products for at least 10 h. On arrival, fasted blood and urine samples were collected and processed for analysis, including hematology, chemistries, liver function tests, thyroid function tests, and urinalysis. Vital signs (heart rate, seated and standing blood pressure), and body anthropometrics (height, weight, waist circumference) were assessed after the participant had been at rest. Cognitive and physical function were assessd, and quality of life questionnaires were administered during Visits 3 and 5; as these results will be described separately, detailed procedures are not described here, but can be found in the detailed description of the protocol [36].

### 2.6 At-Home Procedures

Each day, at home, participants consumed their regular first meal of the day and consumed the study beverage within 5 minutes of finishing their meal. Participants were asked to complete the BTQ 3-6 hours after consumption of the study beverage.

### 2.7 Study Outcomes

#### 2.7.1 Primary outcome

The primary outcome of the BIKE Study was tolerability of the KE intervention, assessed using the BTQ. Specifically, the prespecified primary outcome was the proportion of participants reporting the same moderate to severe symptom (among dizziness, headache, or nausea) occurring on more than one day within any given two week recall period (after week 0–2 acclimation period) during a phone interview. Headache, nausea, and dizziness were selected as they occurred significantly more often at mild severity in our previous 28-day study of a KE in healthy younger adults [34]. The ‘moderate to severe’ intensity was selected as ‘mild’ symptoms are common in this population [37], and may be acceptable if they are outweighed by benefits of the intervention. Moderate to severe symptoms are less common 1. [38] and more likely to be detrimental to quality of life or function. The frequency of ‘more than one day per period’ is used to reduce the impact of idiosyncratic symptoms on the analysis given the common occurrence of such symptoms in the target population.

#### 2.7.2 Secondary outcomes

The secondary outcome of the BIKE Study was safety, assessed through changes over time in clinical and laboratory measures throughout the study, within each group. Changes in body weight, waist circumference, heart rate, systolic and diastolic blood pressure, and orthostatic changes in blood pressure were followed at the last three study visits. The key safety outcome is the within-group change in safety blood parameters. The occurrence of adverse events was also tabulated.

### 2.8 Biological Sample Analysis

All blood and urine sample analysis for clinical laboratory measures took place at accredited Quest Diagnostics laboratories in California. The clinical chemistry profile included albumin, alkaline phosphatase, alanine transaminase (ALT), aspartate aminotransferase (AST), total bilirubin, blood urea nitrogen, creatinine with glomerular filtration rate, carbon dioxide, chloride, globulin, glucose, potassium, total protein, and sodium. A standard lipid panel was performed, including total cholesterol, high density lipoprotein (HDL), low density lipoprotein (LDL), non HDL cholesterol, and triglycerides. High-sensitivity C-reactive protein and a thyroid panel, including triiodothyronine uptake, total thyroxine (T4), and free T4 index were measured. The hematology profile included white blood cell count, red blood cell count, hemoglobin, hematocrit, mean corpuscular volume, mean corpuscular hemoglobin concentration, neutrophils, lymphocytes, monocytes, eosinophils, basophils, and platelet count. Urinalysis parameters included glucose, urobilinogen, ketone body, protein, blood cells, bilirubin, specific gravity, and pH. Normal ranges for all values were provided by the analytical lab and accounted for age and gender.

### 2.9 Statistical Methods

All analyses were based on intention to treat and were conducted by a blinded external study statistician. The proportion of participants reporting at least 2 occurrences of the target symptoms in the phone interview during the post acclimation period (week 3 onward) was estimated along with an exact 90% confidence interval (CI) within each group and compared between groups with the Fisher’s exact test. Additionally, the 10-item composite score, defined as the sum of the respective items, was analyzed with a repeated measures model with an unstructured covariance structured imposed upon the residuals. The frequency of each individual symptom was described with counts and percentages overall and by symptom severity for the 14-day acclimiation period (using the daily logs) and in the post acclimication period (using the phone interview). During the 14-day acclimation period, due to the number of no symptom resposnes, the 3-item (dizziness, headache nausea, maximum score 63) and 10-item (maximum score 210) weekly composite score was calculated as the sum of the individual items each day and compared between group with the Wilcoxon rank sum test. Additionally, the within-group change in safety labs, from baseline to each of the interim and final timepoints, was compared between groups with a Wilcoxon rank sum test and a false discovery rate adjustment to control for multiple comparisons..

The intent-to-treat (ITT) population comprised data for all participants who were randomized and consumed one serving of the study beverage and was the primary population used to assess safety and tolerability. Additionally, the population of completers was analyzed. Missing data was not imputed.

## 3.0 Results

### 3.1 Participants and completion

Participant anthropometric characteristics at baseline are shown in **Table 1**. A total of 30 participants were randomized; 1 participant completed the acute kinetics visit but did not start the main study without giving a reason, and 6 participants dropped out after Day 0 and did not complete the full protocol (KE n= 2 (2F), PLA n = 4 (3F, 1M)). The reasons and times of withdrawal were as follows: GI issues (KE n = 2, week 3 and 5, one participant self-withdrew, one participant withdrawn by medical officer; PLA n =1, week 4, participant withdrew), tiredness (PLA n = 2, week 2 and 6, both participants self-withdrew), and increased cholesterol in safety labs (PLA n = 1, week 4, withdrawn by medical officer). The analysis population (n = 29) consisted of all participants who started the main study, a CONSORT diagram is shown in **Supplemental Figure 1.**

**Table 1:**
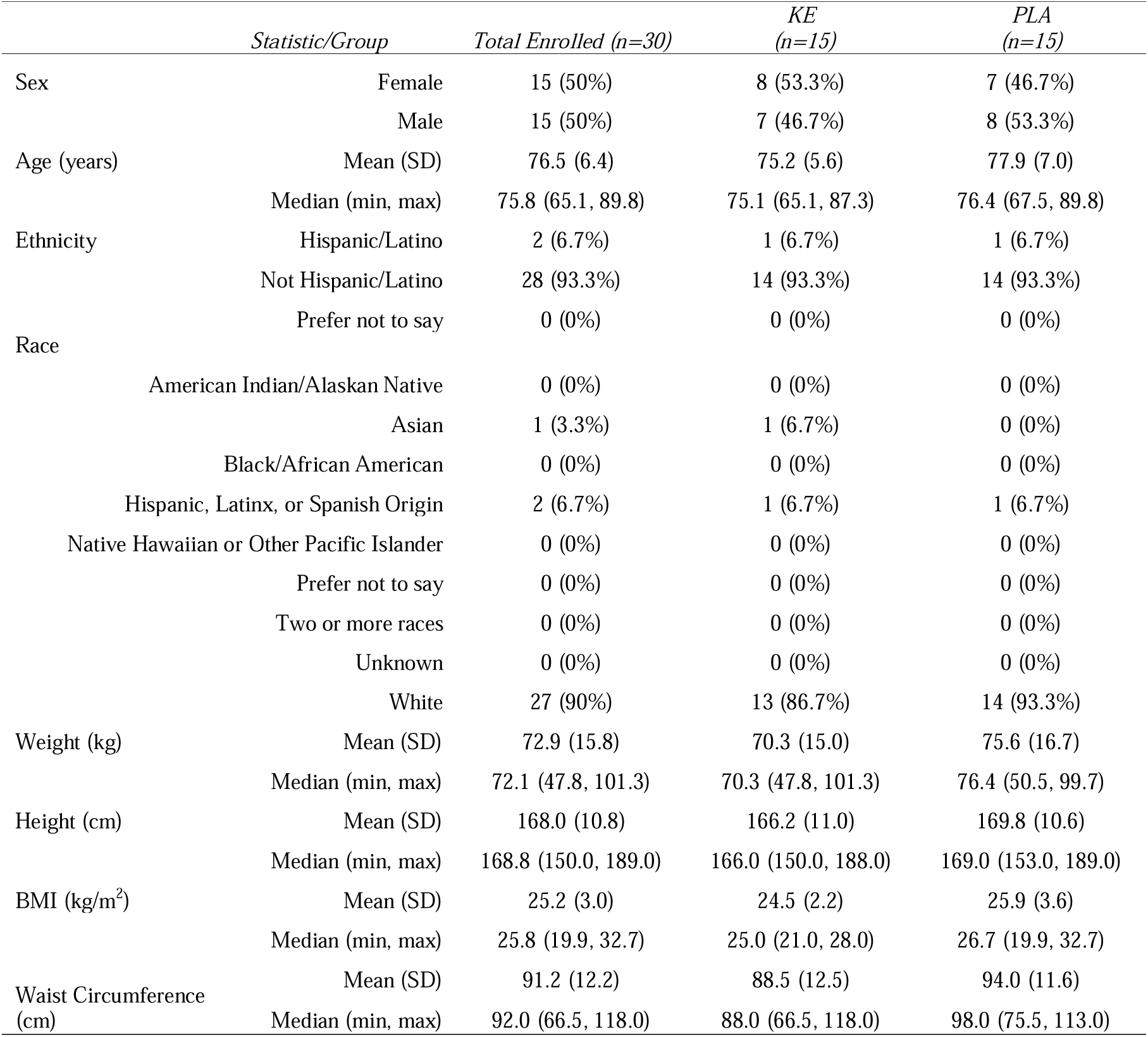
Study participant characteristics. Description of characteristics of randomized study participants. Data represents all individuals’ values obtained at their screening visit. **Abbreviations**: BMI, body mass index; KE, ketone ester; max, maximum; min, minimum; PLA, placebo; SD, standard deviations.

|  | *Statistic/Group* | *Total Enrolled (n=30)* | *KE*<br>*(n=15)* | *PLA*<br>*(n=15)* |
| --- | --- | --- | --- | --- |
| Sex | Female | 15 (50%) | 8 (53.3%) | 7 (46.7%) |
|  | Male | 15 (50%) | 7 (46.7%) | 8 (53.3%) |
| Age (years) | Mean (SD) | 76.5 (6.4) | 75.2 (5.6) | 77.9 (7.0) |
|  | Median (min, max) | 75.8 (65.1, 89.8) | 75.1 (65.1, 87.3) | 76.4 (67.5, 89.8) |
| Ethnicity | Hispanic/Latino | 2 (6.7%) | 1 (6.7%) | 1 (6.7%) |
|  | Not Hispanic/Latino | 28 (93.3%) | 14 (93.3%) | 14 (93.3%) |
|  | Prefer not to say | 0 (0%) | 0 (0%) | 0 (0%) |
| Race | American Indian/Alaskan Native | 0 (0%) | 0 (0%) | 0 (0%) |
|  | Asian | 1 (3.3%) | 1 (6.7%) | 0 (0%) |
|  | Black/African American | 0 (0%) | 0 (0%) | 0 (0%) |
|  | Hispanic, Latinx, or Spanish Origin | 2 (6.7%) | 1 (6.7%) | 1 (6.7%) |
|  | Native Hawaiian or Other Pacific Islander | 0 (0%) | 0 (0%) | 0 (0%) |
|  | Prefer not to say | 0 (0%) | 0 (0%) | 0 (0%) |
|  | Two or more races | 0 (0%) | 0 (0%) | 0 (0%) |
|  | Unknown | 0 (0%) | 0 (0%) | 0 (0%) |
|  | White | 27 (90%) | 13 (86.7%) | 14 (93.3%) |
| Weight (kg) | Mean (SD) | 72.9 (15.8) | 70.3 (15.0) | 75.6 (16.7) |
|  | Median (min, max) | 72.1 (47.8, 101.3) | 70.3 (47.8, 101.3) | 76.4 (50.5, 99.7) |
| Height (cm) | Mean (SD) | 168.0 (10.8) | 166.2 (11.0) | 169.8 (10.6) |
|  | Median (min, max) | 168.8 (150.0, 189.0) | 166.0 (150.0, 188.0) | 169.0 (153.0, 189.0) |
| BMI (kg/m<sup>2</sup>) | Mean (SD) | 25.2 (3.0) | 24.5 (2.2) | 25.9 (3.6) |
|  | Median (min, max) | 25.8 (19.9, 32.7) | 25.0 (21.0, 28.0) | 26.7 (19.9, 32.7) |
| Waist Circumference (cm) | Mean (SD) | 91.2 (12.2) | 88.5 (12.5) | 94.0 (11.6) |
|  | Median (min, max) | 92.0 (66.5, 118.0) | 88.0 (66.5, 118.0) | 98.0 (75.5, 113.0) |

Adherance with consumption of the study products for the overall population was high, with n = 15 reporting 100% adherance and a range of 94 – 99 % for the remaining 8 participants who completed the study, based on study logs and returned bottles.

### 3.2 Tolerance

There was no evidence to support that there was a difference between groups in the study primary end point: the proportion of participants experiencing moderate or severe dizziness, nausea, or headache on more than one day during any of the two-week post-acclimation time periods (**Table 2**). Similarly, there was no evidence of a difference between groups in the proportion of participants reporting any of the 10 items at moderate or severe intensity on more than one day in any of the two-week post acclimation periods.

**Table 2:**
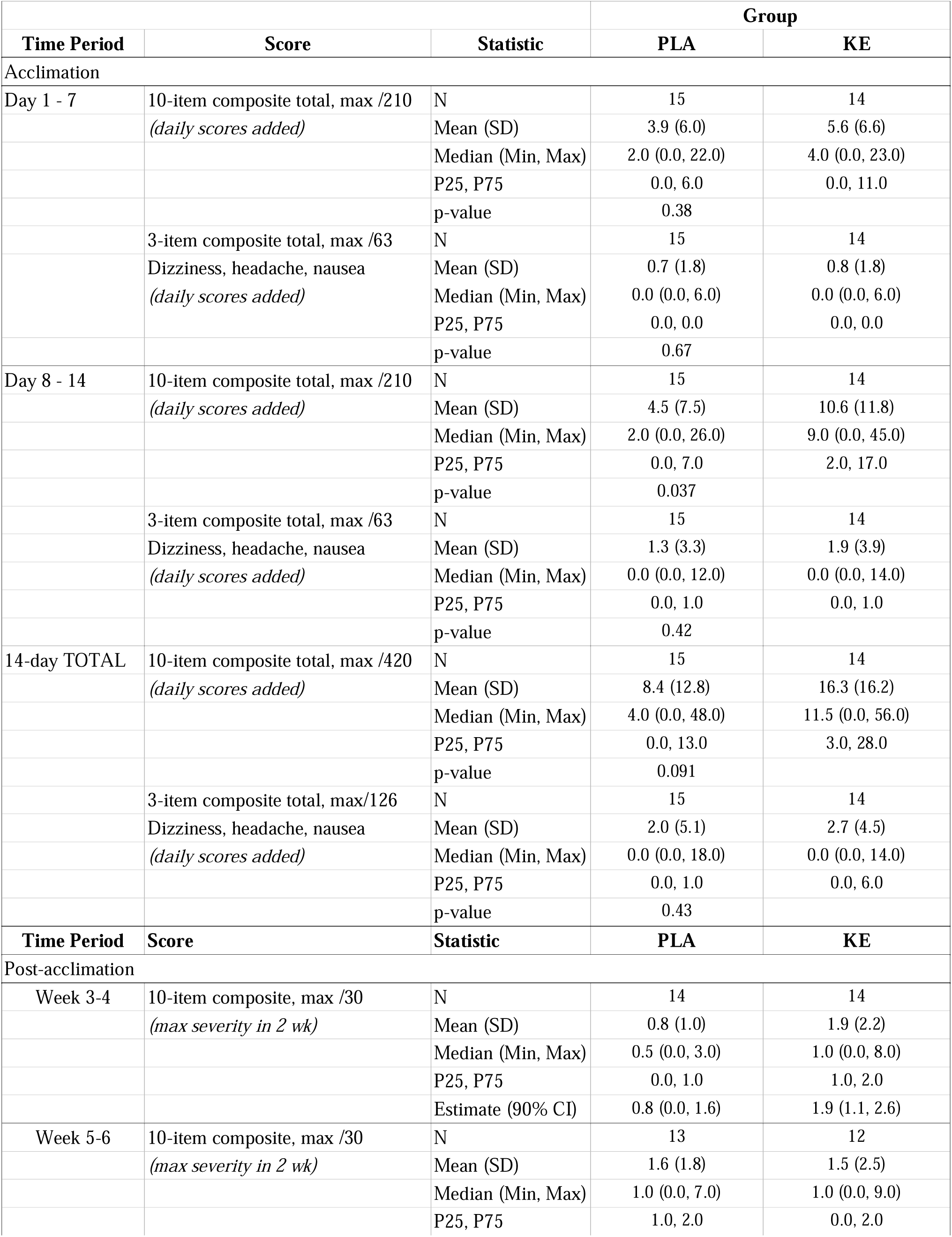
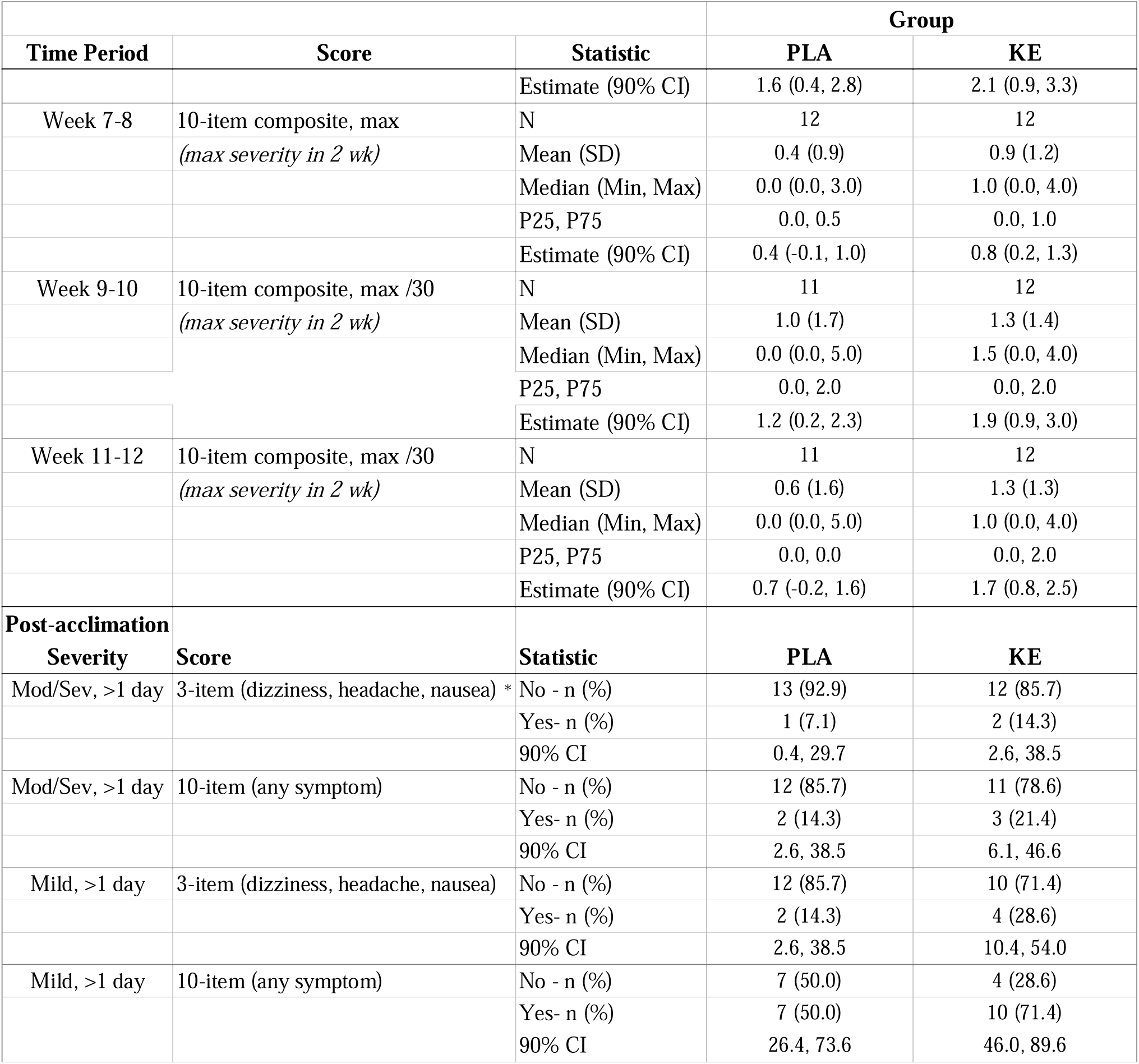
Tolerance outcomes. Tolerance data for healthy older adults consuming ketone ester or placebo daily for 12 weeks. Data is shown for the acclimation period, where serving was escalated from 12.5 to 25 g daily (assessed by daily questionnaire), and for the 10 weeks post acclimation (assessed in bi-weekly phone interview). Data is shown for all 10 items on the beverage tolerance questionnaire as well as the 3 pre-selected items of interest (dizziness, nausea, headache).Proportion of participants with mild OR moderate/severe symptoms on more than one day is show; the primary study outcome is indicated with *. **Abbreviations:** CI, confidence interval; Mod, moderate; Sev, severe, PLA, placebo; KE, ketone ester. All data are ‘all reported’ at each time point.

|  |  |  | Group |  |
| --- | --- | --- | --- | --- |
| Time Period | Score | Statistic | PLA | KE |
| Acclimation |  |  |  |  |
| Day 1 - 7 | 10-item composite total, max /210 | N | 15 | 14 |
|  | (daily scores added) | Mean (SD) | 3.9 (6.0) | 5.6 (6.6) |
|  |  | Median (Min, Max) | 2.0 (0.0, 22.0) | 4.0 (0.0, 23.0) |
|  |  | P25, P75 | 0.0, 6.0 | 0.0, 11.0 |
|  |  | p-value | 0.38 |  |
|  | 3-item composite total, max /63 | N | 15 | 14 |
|  | Dizziness, headache, nausea | Mean (SD) | 0.7 (1.8) | 0.8 (1.8) |
|  | (daily scores added) | Median (Min, Max) | 0.0 (0.0, 6.0) | 0.0 (0.0, 6.0) |
|  |  | P25, P75 | 0.0, 0.0 | 0.0, 0.0 |
|  |  | p-value | 0.67 |  |
| Day 8 - 14 | 10-item composite total, max /210 | N | 15 | 14 |
|  | (daily scores added) | Mean (SD) | 4.5 (7.5) | 10.6 (11.8) |
|  |  | Median (Min, Max) | 2.0 (0.0, 26.0) | 9.0 (0.0, 45.0) |
|  |  | P25, P75 | 0.0, 7.0 | 2.0, 17.0 |
|  |  | p-value | 0.037 |  |
|  | 3-item composite total, max /63 | N | 15 | 14 |
|  | Dizziness, headache, nausea | Mean (SD) | 1.3 (3.3) | 1.9 (3.9) |
|  | (daily scores added) | Median (Min, Max) | 0.0 (0.0, 12.0) | 0.0 (0.0, 14.0) |
|  |  | P25, P75 | 0.0, 1.0 | 0.0, 1.0 |
|  |  | p-value | 0.42 |  |
| 14-day TOTAL | 10-item composite total, max /420 | N | 15 | 14 |
|  | (daily scores added) | Mean (SD) | 8.4 (12.8) | 16.3 (16.2) |
|  |  | Median (Min, Max) | 4.0 (0.0, 48.0) | 11.5 (0.0, 56.0) |
|  |  | P25, P75 | 0.0, 13.0 | 3.0, 28.0 |
|  |  | p-value | 0.091 |  |
|  | 3-item composite total, max/126 | N | 15 | 14 |
|  | Dizziness, headache, nausea | Mean (SD) | 2.0 (5.1) | 2.7 (4.5) |
|  | (daily scores added) | Median (Min, Max) | 0.0 (0.0, 18.0) | 0.0 (0.0, 14.0) |
|  |  | P25, P75 | 0.0, 1.0 | 0.0, 6.0 |
|  |  | p-value | 0.43 |  |
| Time Period | Score | Statistic | PLA | KE |
| Post-acclimation |  |  |  |  |
| Week 3-4 | 10-item composite, max /30 | N | 14 | 14 |
|  | (max severity in 2 wk) | Mean (SD) | 0.8 (1.0) | 1.9 (2.2) |
|  |  | Median (Min, Max) | 0.5 (0.0, 3.0) | 1.0 (0.0, 8.0) |
|  |  | P25, P75 | 0.0, 1.0 | 1.0, 2.0 |
|  |  | Estimate (90% CI) | 0.8 (0.0, 1.6) | 1.9 (1.1, 2.6) |
| Week 5-6 | 10-item composite, max /30 | N | 13 | 12 |
|  | (max severity in 2 wk) | Mean (SD) | 1.6 (1.8) | 1.5 (2.5) |
|  |  | Median (Min, Max) | 1.0 (0.0, 7.0) | 1.0 (0.0, 9.0) |
|  |  | P25, P75 | 1.0, 2.0 | 0.0, 2.0 |
| Time Period | Score | Statistic | PLA | KE |
|  |  | Estimate (90% CI) | 1.6 (0.4, 2.8) | 2.1 (0.9, 3.3) |
| Week 7-8 | 10-item composite, max | N | 12 | 12 |
|  | (max severity in 2 wk) | Mean (SD) | 0.4 (0.9) | 0.9 (1.2) |
|  |  | Median (Min, Max) | 0.0 (0.0, 3.0) | 1.0 (0.0, 4.0) |
|  |  | P25, P75 | 0.0, 0.5 | 0.0, 1.0 |
|  |  | Estimate (90% CI) | 0.4 (-0.1, 1.0) | 0.8 (0.2, 1.3) |
| Week 9-10 | 10-item composite, max /30 | N | 11 | 12 |
|  | (max severity in 2 wk) | Mean (SD) | 1.0 (1.7) | 1.3 (1.4) |
|  |  | Median (Min, Max) | 0.0 (0.0, 5.0) | 1.5 (0.0, 4.0) |
|  |  | P25, P75 | 0.0, 2.0 | 0.0, 2.0 |
|  |  | Estimate (90% CI) | 1.2 (0.2, 2.3) | 1.9 (0.9, 3.0) |
| Week 11-12 | 10-item composite, max /30 | N | 11 | 12 |
|  | (max severity in 2 wk) | Mean (SD) | 0.6 (1.6) | 1.3 (1.3) |
|  |  | Median (Min, Max) | 0.0 (0.0, 5.0) | 1.0 (0.0, 4.0) |
|  |  | P25, P75 | 0.0, 0.0 | 0.0, 2.0 |
|  |  | Estimate (90% CI) | 0.7 (-0.2, 1.6) | 1.7 (0.8, 2.5) |
| **Post-acclimation** |  |  |  |  |
| **Severity** | **Score** | **Statistic** | **PLA** | **KE** |
| Mod/Sev, >1 day | 3-item (dizziness, headache, nausea) * | No - n (%) | 13 (92.9) | 12 (85.7) |
|  |  | Yes- n (%) | 1 (7.1) | 2 (14.3) |
|  |  | 90% CI | 0.4, 29.7 | 2.6, 38.5 |
| Mod/Sev, >1 day | 10-item (any symptom) | No - n (%) | 12 (85.7) | 11 (78.6) |
|  |  | Yes- n (%) | 2 (14.3) | 3 (21.4) |
|  |  | 90% CI | 2.6, 38.5 | 6.1, 46.6 |
| Mild, >1 day | 3-item (dizziness, headache, nausea) | No - n (%) | 12 (85.7) | 10 (71.4) |
|  |  | Yes- n (%) | 2 (14.3) | 4 (28.6) |
|  |  | 90% CI | 2.6, 38.5 | 10.4, 54.0 |
| Mild, >1 day | 10-item (any symptom) | No - n (%) | 7 (50.0) | 4 (28.6) |
|  |  | Yes- n (%) | 7 (50.0) | 10 (71.4) |
|  |  | 90% CI | 26.4, 73.6 | 46.0, 89.6 |

Other tolerance analyses were exploratory in nature and focused on temporal changes through the study, and on other symptom types and severity. The first two weeks of the intervention was an acclimation period, where serving size increased from 12.5 to 25 g from Day 8 onwards, and participants completed a tolerance questionnaire every day. During this acclimation period n = 5 participants (KE = 1, PLA = 4) experienced no symptoms at any time, n = 8 participants reported at least one ‘mild’ symptom (KE =5, PLA = 3), n = 13 participants reported at least one ‘moderate’ symptoms (KE = 6, PLA = 7), and n =3 participants experienced one ‘severe’ symptoms (KE = 2, dizziness on day 6, dizziness on day 14; PLA = 1, headache on day 7).

As there was a high rate of ‘no’ symptom reports each day for both groups, three (dizziness, headache and nausea) and ten item composite scores for the tolerance questionnaire were calculated as the total of all scores on days 1-7 (week 1 composite) and 8 – 14 (week 2 composite) (**Table 2)**. There was no difference between groups in the week 1 composite for the three major symptom items (p = 0.67), or in the week 1 composite score for all ten symptom items (p = 0.38, **Figure 2A**). In the second week, when study product serving size increased, there was no difference in the week 2 composite score for the three major items (p = 0.42), but there was a significantly greater week 2 composite score for all ten items in the KE group compared with the PLA group (p = 0.037, **Figure 3A**). When the 14-day acclimation period was considered overall, (day 1 -14 composite), there was no difference in the three-item composite score (p = 0.43, **Table 2**). However, a significant difference was suggested in the distribution of the ten item composite, where the KE group tended higher (median (25^th^, 75^th^ percentile (KE = 11.5 (3, 28); PLA = 4 (0, 13) p =0.091).

**Figure 2:**
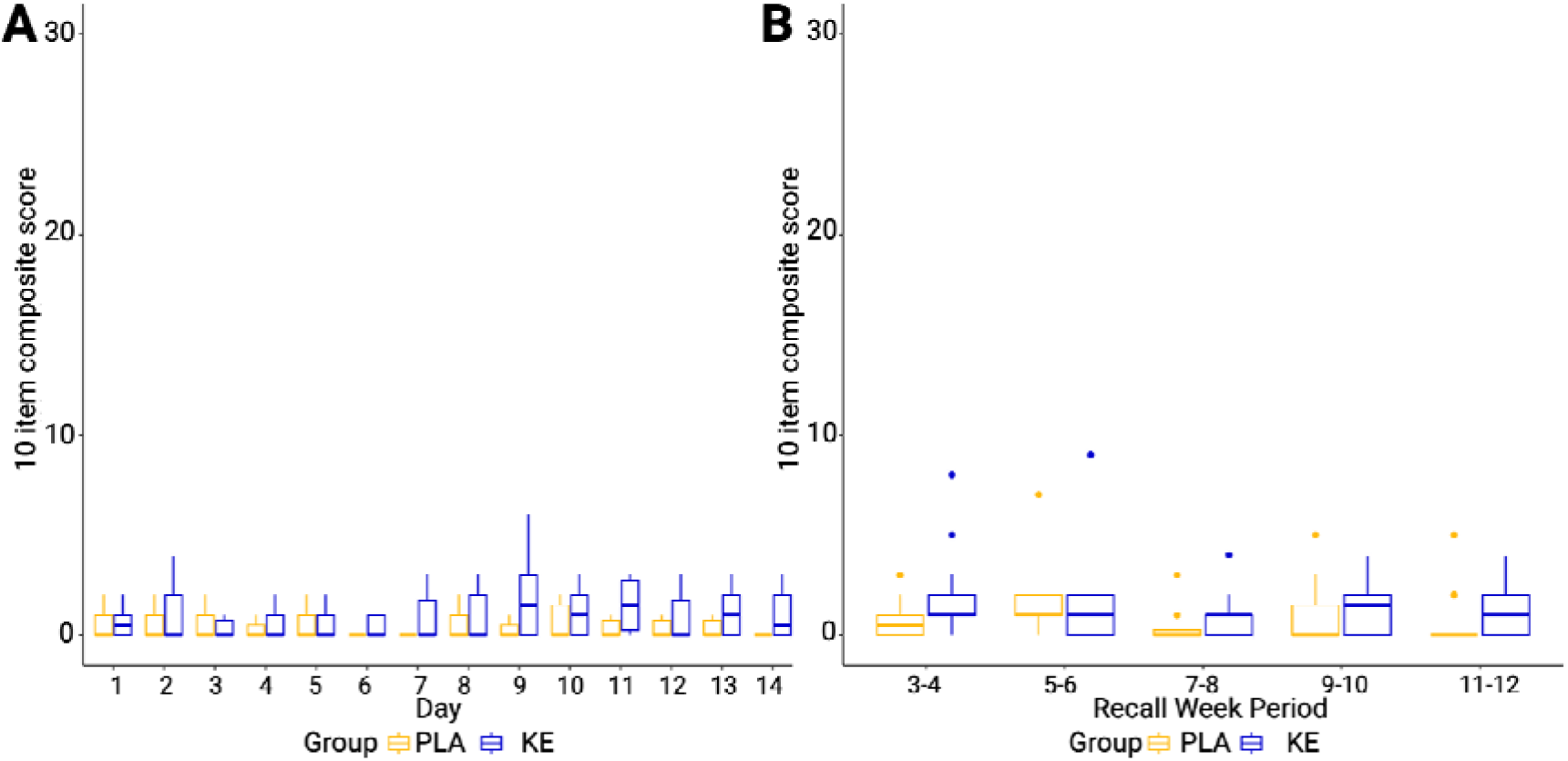
Composite scores for all 10 items assesed in the Bevereage Tolerability Questionnaire (BTQ) either by daily duirng questionnaire during the two week acclimation period (**A)** or by bi-weekly phone interview in the post acclimation period (**B).** The maximum score is 30.

In the post-acclimation period between 3 – 12 weeks (**Table 2**) participants continued to consume the 25 g serving size, but tolerance was assessed less frequently - every 2 weeks – by interview and referral to the daily study log where symptoms could be recorded. There was no main effect of group on the ten-item tolerance score (p = 0.20, **Figure 2B**), a composite of all reported symptoms at their maximum severity, and no group x time interaction (p = 0.89), or time (p = 0.36). No post hoc testing was performed due to the absence of an interaction effect.

In parallel to our primary outcome which focused on moderate to severe symptoms, we also analyzed the proportion of participants who experienced mild dizziness, nausea, or headache at least twice in a two-week period, post acclimation. This was not different between groups. The proportion of participants who reported any of the ten items at least twice at a mild intensity in a two-week period, post acclimation, was also not different between groups.

When all symptoms were considered, the most common symptoms reported at least once during acclimation were abdominal cramping (n = 5) and burping (n = 5), and post acclimation were abdominal cramping (n = 6) and reflux (n = 6) for the KE group.

Overall, it appeared that up to 25 g of KE taken daily did not meaningfully increase the occurrence of either the expected symptoms based on younger adult studies (dizziness, nausea, or headache), or other symptoms not previously linked to KE consumption.

### 3.3 Safety

Consumption of up to 25 g per day of KE or PLA by older adults was not associated with any statistically significant or clinically relevant changes in vital signs (**Table 3**) or any of the laboratory tests conducted: clinical chemistry, hematology, lipid panel, thyroid panel, or urinalysis; selected laboratory values that were expected to be affected by KE metabolism are shown in **Table 3**; all were unchanged. As described above, two participants dropped out of the KE group due to GI issues, compared to four participants dropping out of the placebo group; all dropouts were judged to be at least ‘possibly’ related to the study products. There were no study product related AEs reported during the study. Seven participants in total reported non-related AEs including: twisted ankle, upper respiratory tract infection (n = 3), broken toe, hairline foot fracture, and back pain.

**Table 3:**
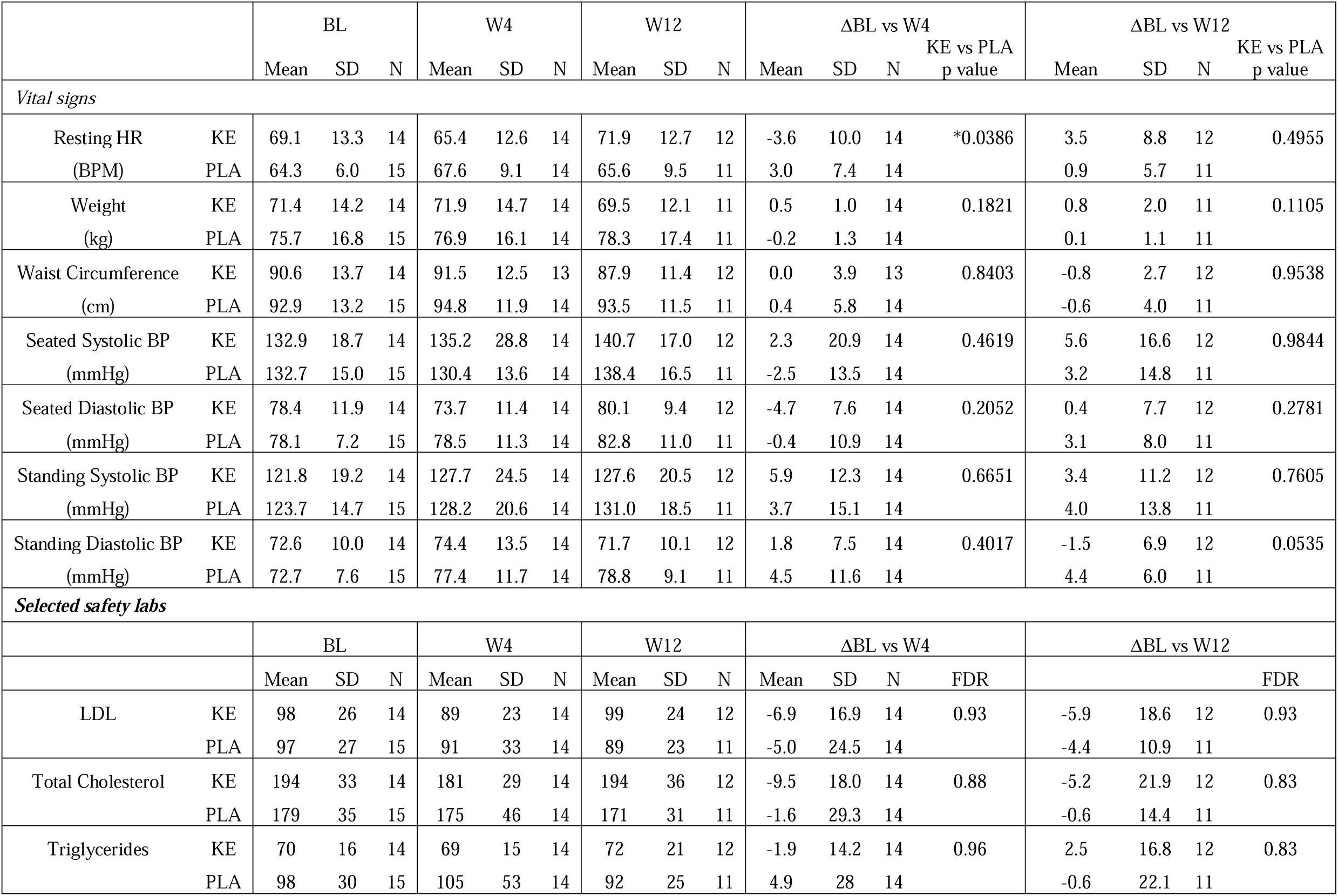
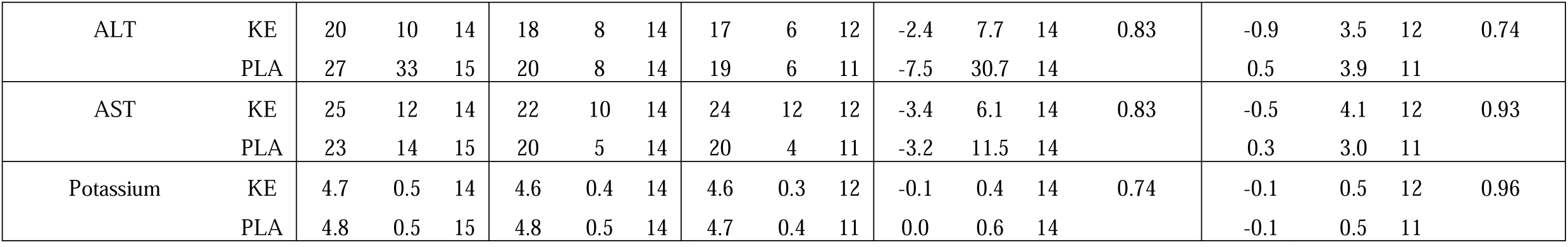
Vital signs and selected safety labs. Vital signs, morphological data and selected safety labs (based on expected interaction with ketone biology) from older adults consuming KE or A at baseline, week 4 visit and week 12 visit, and delta from baseline at both week 4 and week 12. P value is for KE vs PLA, * indicates p < 0.05.. **Abbreviations**: BL, baseline; BP, blood pressure; KE, ketone ester; PLA, placebo; HR, heart rate; BPM, beats per minute.

## 4.0 Discussion

The results of this study demonstrate that healthy older adults can tolerably and safely consume up to 25 g per day of the ketone ester, bis-octanoyl-(R)1-3, butanediol. These findings considerably extend the duration of previous KE tolerance and safety studies, as well as provide the first data exclusively in older adults. This establishes a critical platform for future studies investigating exogenous ketones and ketone esters as a geroscience intervention, addressing age-related disease by targeting aging biology.

Despite previous studies of KE finding good adherence, limited symptoms, and no safety concerns, exogenous ketones have a reputation for being difficult to implement in long term interventions [41]. Acute side-effects of a similar nature to those seen in this study (i.e., nausea, headache, dizziness) have been seen with all types of exogenous ketones products that are functionally similar to BO-BD, including medium chain triglycerides (MCT), 1,3-butanediol, ketone salts, acetoacetate ketone di-ester, and BHB-ketone mono-ester [42–45]. These side-effects are seen at a range of serving sizes (10 - 85g), under different conditions (fed, fasted, rest, exercise), and manifest as both acute GI discomfort and distress, as well as systemic symptoms such as headache, nausea, and dizziness that range in frequency and intensity from mild to severe. In the current study, symptoms occurred at a similar rate in both ketone ester and placebo groups, and symptoms were not more frequent than seen in studies of BH-BD or BO-BD in younger adults [11, 34], despite the expectation that older adults might have either an increased risk of intervention-related symptoms or an increased rate of non-related, sporadic symptoms. Overall, these data support the conclusion that BO-BD is well tolerated in healthy older adults at the serving sizes studied.

Alongside tolerance, an often cited concern with longer term studies of ketone esters is lack of adherence due to poor palatability [41], leading investigators to compare product palatability as an important component of intervention selection [46]. In a recent study, the sensory characteristics of BO-BD to BH-BD were compared and found BO-BD was more palatable [11]. In the current study, the high adherence rate of participants indicates that this KE was acceptable, and that palatability issues would not be a barrier to longer term implementation.

A final area of concern is safety of long-term KE consumption, which was a key endpoint of this study given the expected vulnerability of the older adult population. Based on previous studies of exogenous ketones, there were several areas that were carefully monitored here. Firstly, as KE have previously been seen to suppress appetite [47], which would be undesirable in an older population, we monitored body weight and saw no change in either group. Secondly, as the KE is a lipid-based molecule that is metabolized in the liver and the canola oil placebo contained lipids, we closely tracked changes to lipid panels (withdrawing one participant in the PLA group) and liver function tests. No changes in these lab tests were deemed clinically significant by the independent study medical officers. Thirdly, exogenous ketones can acutely alter blood acid-base balance and electrolytes [48] which could be problematic either in isolation or via medication interactions; no changes were seen in related endpoints here. Finally, we monitored vital signs, as exogenous ketones have been demonstrated to acutely increase heart rate and decrease blood pressure [49]; these were again unchanged in this study. Notably, we did not see changes in fasting glucose in either group; a decrease in glucose might have been expected in the KE group given the consistent hypoglycemic effect of exogenous ketones [19]. Overall, in this relatively healthy population of older adults, 25 g of KE daily for 12 weeks did not lead to any safety concerns. Still, safety monitoring should remain a key feature of future geroscience studies using ketone esters, as less healthy populations or populations with specific comorbidities may be more vulnerable to metabolic and physiologic perturbation.

Given that there were no tolerance, safety, or compliance concerns raised by this study, increased KE dosing could be considered in future work. Up to 50 g/day for one week of BO-BD per day has been studied in younger adults [11], and servings of up to 75 g /day for 28 days of other KE has been reported [32, 33]. As we saw symptoms increase with increasing dose between days 8 – 14 and then decline over the remainder of the study, we suggest that gradually increasing the daily serving over several weeks may provide optimal tolerance. This would be consistent with the gradually increasing serving size recommended in clinical protocols for MCT use in therapeutic ketogenic diets [50]. The advantage of providing a higher dose, split into multiple servings across the day, would be an increased time in a state of hyper-ketonemia. It is currently unknown if greater peak ketone concentrations or greater time of ketone exposure time drives clinical efficacy. Studies that have found benefits of exogenous ketones in metabolic [19], cardiac [49], and cognitive [51] function have linked blood ketone concentration to improved efficacy. These observations reinforce the need to use the maximal tolerable and safe dose in studies designed to explore new applications of nutritional ketosis.

The strengths of this study include the free-living, pragmatic-inspired design (e.g. participants were instructed to maintain their usual diet and exercise habits) being highly relevant for future uses of KE as a geroscience intervention, the high adherence observed, the equal enrollment of men and women, the average age (76 y) being well over the lower limit (65 y), and the close matching achieved between the KE and PLA beverage. The weaknesses of this study were to be expected, given that it was planned as a pilot to help design a larger clinical trial targeting physical function related to frailty in older adults. Firstly, the sample size was small, and predominantly identified as non-Hispanic white; results in this population may not be generalizable to larger studies or to all populations of older adults. Secondly, the overall incidence of dropouts was 23%, slightly higher than our expected 20%, but this rate decreases if accounting for the participant who was randomized but did not start the 12-week study, and more participants in the PLA group did not complete the study compared to the KE group. Thirdly, we relied on beverage logs and bottle return to confirm compliance but there is not a validated biomarker that allows us to verify that beverage consumption was accurately reported. Finally, although we encouraged participants to use the beverage log to record any symptoms in their daily log, they did not have to complete a full questionnaire every day after day 14, meaning some symptoms may have been forgotten and not reported. We expect that this effect might be more relevant to mild symptoms, as moderate and severe symptoms would be more disruptive and more likely to be remembered and reported.

In conclusion this pilot study demonstrated the safety and tolerability of 12-weeks of consumption of up to 25 g per day of the ketone di-ester, bis-octanoyl-(R)-1,3-butanediol in healthy older adults. These findings will facilitate the application of ketone body biology in human health and disease in aging.

## Author Declarations and Conflict Management

The Buck Institute holds shares in BHB Therapeutics (Ireland) and Selah Therapeutics. B.J.S. has stock in H.V.M.N Inc, and stock options in Selah Therapeutics Ltd, BHB Therapeutics (Ireland) Ltd., and Juvenescence Ltd. J.C.N. has stock options in Selah Therapeutics Ltd and BHB Therapeutics (Ireland) Ltd. J.C.N and B.J.S. are inventors on patents related to the use of ketone bodies that are assigned to The Buck Institute. Individual and institutional conflict management plans were developed and approved by the Buck Institute and submitted to the reviewing IRB. Actions and decisions important to participant safety and study integrity were carried out by ‘honest brokers’ with no potential financial conflict. Participant consent was obtained by licensed registered nurses (L.A and W.S.M) who have no financial conflict. Decisions on participant enrollment, continuation, and were made by independent medical officers (J.M and M.Y) unaffiliated with Buck Institute and with no financial conflict. Data analysis for the primary outcome was carried out by an independent statistician with no financial conflict (T.B). All other authors have no conflicts to report.

## Author Contributions

Conceptualization, J.C.N, B.J.S; methodology, J.C.N, B.J.S; investigation, B.J.S, E.B.S, C.S, S.R.D, S.P, W.S-M, L.A, M.Y, J.M; data curation, B.J.S, E.B.S, C.S, S.R.D, S.P; formal analysis, T.B, B.J.S; writing—original draft preparation, B.J.S, E.B.S; writing—review and editing, all; visualization, B.J.S, T.B; project administration, B.J.S, J.C.N, T.G; supervision: B.J.S, T.G, J.C.N; funding acquisition, B.J.S, J.C.N; All authors have read and agreed to the published version of the manuscript.

## Data Availability Statement

The data presented here may be available upon reasonable request from the corresponding author and in accordance with intellectual property considerations.

## Supporting information

CONSORT Checklist

## Abbreviations

ALT: Alanine Transaminase
AST: Aspartate Aminotransferase
BH-BD: Bis-hexanoyl (R)-1,3-butanediol
BHB: Beta-hydroxybutyrate
BIKE: Buck Institute Ketone Ester
BMI: Body Mass Index
BO-BD: Bis-octanoyl (R)-1,3-butanediol
BTQ: Beverage Tolerability Questionnaire
CI: Confidence Interval
GI: Gastrointestinal
HDL: High Density Lipoprotein
ITT: Intent to Treat
KE(s): Ketone Ester(s)
LDL: Low Density Lipoprotein
MCT: Medium Chain Triglyceride
PLA: Placebo
PP: Per Protocol
T4: Thyroxine

---

**Supplementary Table 1.** Nutritional Information. Nutritional information for the KE and PLA beverages. **Abbreviations:** BO-BD, bis-octanoyl (R)-1,3 butanediol; KE, ketone ester; PLA, placebo

|  | KE | PLA |
| --- | --- | --- |
| BO-BD (g) | 25 | 0 |
| Energy (kcal) | 240 | 246 |
| Carbohydrate (g) | 2 | 2 |
| Fat (g) | 0.5 | 25 |
| Protein (g) | 2 | 2 |

**Supplementary Table 2.** Symptom reports during bi-weekly phone interview, listed by type and severity. Number of participants that reported during a bi-weekly phone interview that the listed symptoms occurred at least once during the pre- (week 1 and 2) and post- (week 3 – 12) acclimation phases of the study. **Abbreviations:** KE, ketone ester.

| Symptom | Severity | Acclimation Period |  | Post Acclimation Period |  |
| --- | --- | --- | --- | --- | --- |
|  |  | Placebo | KE | Placebo | KE |
| Any | Any | 9 (60.0) | 12 (85.7) | 10 (71.4) | 14 (100.0) |
|  | Mild | 9 (60.0) | 11 (78.6) | 10 (71.4) | 13 (92.9) |
|  | Moderate/Severe | 4 (26.7) | 6 (42.9) | 5 (35.7) | 7 (50.0) |
| Abdominal cramping | Any | 2 (13.3) | 5 (35.7) | 6 (42.9) | 6 (42.9) |
|  | Mild | 1 (6.7) | 4 (28.6) | 5 (35.7) | 5 (35.7) |
|  | Moderate/Severe | 1 (6.7) | 3 (21.4) | 2 (14.3) | 1 (7.1) |
| Burping | Any | 4 (26.7) | 5 (35.7) | 3 (21.4) | 4 (28.6) |
|  | Mild | 4 (26.7) | 5 (35.7) | 3 (21.4) | 4 (28.6) |
|  | Moderate/Severe | 0 (0.0) | 1 (7.1) | 0 (0.0) | 0 (0.0) |
| Diarrhea | Any | 4 (26.7) | 4 (28.6) | 5 (35.7) | 4 (28.6) |
|  | Mild | 4 (26.7) | 3 (21.4) | 5 (35.7) | 3 (21.4) |
|  | Moderate/Severe | 3 (20.0) | 2 (14.3) | 3 (21.4) | 2 (14.3) |
| Dizziness | Any | 1 (6.7) | 1 (7.1) | 1 (7.1) | 6 (42.9) |
|  | Mild | 1 (6.7) | 1 (7.1) | 0 (0.0) | 3 (21.4) |
|  | Moderate/Severe | 1 (6.7) | 1 (7.1) | 1 (7.1) | 3 (21.4) |
| Gas | Any | 4 (26.7) | 5 (35.7) | 5 (35.7) | 4 (28.6) |
|  | Mild | 4 (26.7) | 5 (35.7) | 5 (35.7) | 4 (28.6) |
|  | Moderate/Severe | 1 (6.7) | 2 (14.3) | 2 (14.3) | 1 (7.1) |
| Headache | Any | 1 (6.7) | 2 (14.3) | 2 (14.3) | 4 (28.6) |
|  | Mild | 1 (6.7) | 1 (7.1) | 1 (7.1) | 4 (28.6) |
|  | Moderate/Severe | 1 (6.7) | 1 (7.1) | 1 (7.1) | 1 (7.1) |
| Nausea | Any | 2 (13.3) | 3 (21.4) | 2 (14.3) | 4 (28.6) |
|  | Mild | 2 (13.3) | 2 (14.3) | 2 (14.3) | 3 (21.4) |
|  | Moderate/Severe | 0 (0.0) | 1 (7.1) | 0 (0.0) | 2 (14.3) |
| Reflux | Any | 1 (6.7) | 4 (28.6) | 1 (7.1) | 5 (35.7) |
|  | Mild | 1 (6.7) | 3 (21.4) | 1 (7.1) | 4 (28.6) |
|  | Moderate/Severe | 0 (0.0) | 2 (14.3) | 0 (0.0) | 3 (21.4) |
| Stomach rumbling | Any | 4 (26.7) | 4 (28.6) | 3 (21.4) | 2 (14.3) |
|  | Mild | 3 (20.0) | 4 (28.6) | 3 (21.4) | 2 (14.3) |
|  | Moderate/Severe | 2 (13.3) | 1 (7.1) | 0 (0.0) | 0 (0.0) |
| Vomiting | Any | 0 (0.0) | 0 (0.0) | 0 (0.0) | 0 (0.0) |
|  | Mild | 0 (0.0) | 0 (0.0) | 0 (0.0) | 0 (0.0) |
|  | Moderate/Severe | 0 (0.0) | 0 (0.0) | 0 (0.0) | 0 (0.0) |

**Supplementary Figure 1.**
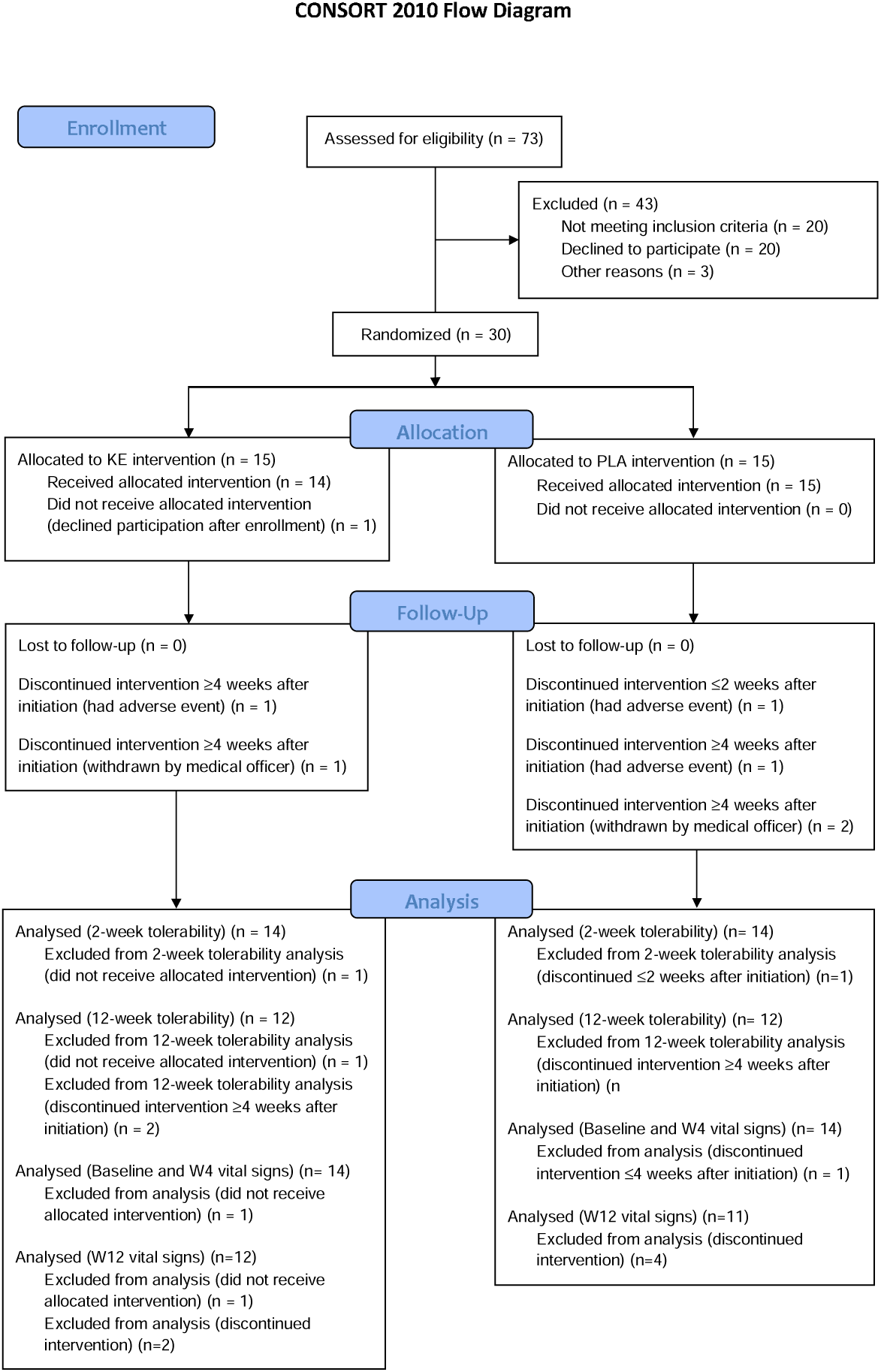

